# Evidence for treatment with estradiol for women with SARS-CoV-2 infection

**DOI:** 10.1101/2020.08.21.20179671

**Authors:** Ute Seeland, Flaminia Coluzzi, Maurizio Simmaco, Cameron Mura, Philip E. Bourne, Max Heiland, Robert Preissner, Saskia Preissner

**Affiliations:** Institute of Physiology, Charité – Universitätsmedizin Berlin, corporate member of Freie Universität Berlin, Humboldt-Universität zu Berlin, and Berlin Institute of Health, Philippstrasse 12, 10115 Berlin; Department Medical and Surgical Sciences and Biotechnologies, Sapienza University of Rome, 04100, Latina, Italy; Unit of Anesthesia and Intensive Care Medicine, Sant’Andrea University Hospital, 00189, Rome, Italy; Department Neurosciences, Mental Health and Sensory Organs, Sapienza University of Rome, 00189, Rome, Italy; Advanced Molecular Diagnostic Unit, Sant’Andrea University Hospital, 00189, Rome, Italy; School of Data Science and Department of Biomedical Engineering, University of Virginia, Charlottesville, VA; USA; Department Oral and Maxillofacial Surgery, Charité *–* Universitätsmedizin Berlin, corporate member of Freie Universität Berlin, Humboldt-Universität zu Berlin, and Berlin Institute of Health Augustenburger Platz 1, 13353 Berlin, Germany

**Keywords:** sex, women, SARS-CoV-2, COVID-19, estradiol, hormone treatment, ACE2

## Abstract

**Background:** Given that an individual’s age and gender are strongly predictive of COVID-19 outcomes, do such factors imply anything about preferable therapeutic options?

**Methods:** An analysis of electronic health records for a large (68,466-case), international COVID-19 cohort, in five-year age strata, revealed age-dependent sex differences. In particular, we surveyed the effects of systemic hormone administration in women. The primary outcome for estradiol therapy was death. Odds Ratios (ORs) and Kaplan-Meier survival curves were analyzed for 37,086 COVID-19 women in two age groups: pre- (15-49 years) and post-menopausal (>50 years).

**Results:** The incidence of SARS-CoV-2 infection is higher in women than men (about +15%) and, in contrast, the fatality rate is higher in men (about +50%). Interestingly, the relationships between these quantities are also linked to age. Pre-adolescent girls had the same risk of infection and fatality rate as boys. Adult premenopausal women had a significantly higher risk of infection than men in the same five-year age stratum (about 16,000 vs. 12,000 cases). This ratio changed again in postmenopausal women, with infection susceptibility converging with men. While fatality rates increased continuously with age for both sexes, at 50 years there was a steeper increase for men. Thus far, these types of intricacies have been largely neglected. Because the hormone 17ß-estradiol has a positive effect on expression of the human ACE2 protein—which plays an essential role for SARS-CoV-2 cellular entry—propensity score matching was performed for the women’s sub-cohort, comparing users versus non-users of estradiol. This retrospective study of hormone therapy in female COVID-19 patients shows that the fatality risk for women >50 yrs receiving estradiol therapy (user group) is reduced by more than 50%; the OR was 0.33, 95 % CI [0.18, 0.62] and the Hazard Ratio was 0.29, 95% CI[0.11,0.76]. For younger, pre-menopausal women (15-49 yrs), the risk of COVID-19 fatality is the same irrespective of estradiol treatment, probably because of higher endogenous estradiol levels.

**Conclusions:** As of this writing, still no effective drug treatment is available for COVID-19; since estradiol shows such a strong improvement regarding fatality in COVID-19, we suggest prospective studies on the potentially more broadly protective roles of this naturally occurring hormone.

## Background

### Epidemiological Data

Early epidemiological observations indicated that SARS-CoV-2 infects all age groups, but with a higher rate among men (58.1%) than women (41.9%) (Guan et al., 2020). The fraction of males among critically ill patients testing positive for COVID-19 is higher, and the outcome is far worse than for women (among casualties, 66% are male and 34% are female) (Yang et al., 2020). From a medical perspective of sex and gender, considering the epidemiological data in a more differentiated manner—based, for instance, on age, sex and lifestyle/behavior patterns—may enable the identification of sub-populations with increased susceptibility (or resistance) to SARS-CoV-2 infection.

As an example of such nuances, consider that spread of the viral infection exhibits a sex-dependence, and also a higher fatality rate amongst older patients, > 60 yrs in age (Verity et al., 2020; Zhou et al., 2020). In addition, smoking behavior has been one of the most appreciated gender differences thus far, and it may underlie the far worse outcomes in men than in women (Cai, 2020); however, this trend has not been confirmed beyond a handful of countries, and the causal aspects of these trends are likely to be rather complex.

### Pathophysiological Context

SARS-CoV-2 virions use the angiotensin-converting enzyme 2 (ACE2) as a host-cell receptor (Hoffmann et al., 2020) for viral uptake. Human ACE2 is an essential part of the renin-angiotensin system (RAS), and is encoded on the X chromosome (Crackower et al., 2002). ACE2 is widely distributed in tissues, including lung alveolar (type II) epithelial cells, the vascular endothelium, heart, kidney, and testis (Gheblawi et al., 2020). It has extensive vascular and organ-protective functions mediated via angiotensin (Ang 1–7) by the angiotensin II receptor type 2 (AT2) and the Mas receptor (MasR). Seeland et al. have reviewed the sexual dimorphism, by virtue of the X chromosomal gene location of ACE2 and the AT2 receptor, as well as the effects of 17ß-estradiol on different components of the renin-angiotensin-aldosterone system (RAAS) (Seeland & Regitz-Zagrosek, 2012). Estrogens downregulate the angiotensin II receptor type 1 (AT1R) signaling pathway by inhibiting angiotensin-converting enzyme (ACE) activity (which, in turn, diminishes the production of angiotensin II peptide). This classical ACE/AngII/AT1R regulatory axis counter-regulates the ACE2/Ang 1–7/MasR axis.

Based on gender-specific differences in genetics and sex hormone levels, the worse outcomes of SARS-CoV-2 infection in men could be triggered by (i) conditions that are associated with a decrease in ACE2 expression, such as older age (Xie, Chen, Wang, Zhang, & Liu, 2006) or(ii) by the more direct loss/depletion of active ACE2 (e.g., because of binding by SARS-CoV-2). Moreover, the regulation of immune cells and cytokine activity are also linked to sex hormone levels. In terms of disease etiology, this could represent another set of factors that underlies the sex differences in pulmonary and vascular symptoms, severity, and outcomes of COVID-19.

## Methods

### Objectives of this Study

As is known from the literature, endogenous estradiol has beneficial cardiovascular and immune system-stabilizing effects in premenopausal women, as well as within pregnancy(Dubey & Jackson, 2001; Straub, 2007). We hypothesize that exogenous estradiol intake is a protective strategy in pre- and post-menopausal women suffering from SARS-CoV-2 infection.

This study assesses the effects of exogenous sex hormone intake from oral contraceptives by premenopausal women, and estradiol hormone therapy by postmenopausal women with SARS-CoV-2 infection or COVID-19 disease on the outcome death.

### Database and Inclusion Criteria

COVID-19 patients were identified via ICD-10 code U07.1 or the presence of SARS-CoV-2-related RNA diagnosis within the last six months. (We did not include diagnoses coded as U07.2 [“COVID-19, virus not identified”] because “laboratory confirmation is inconclusive or not available” in such cases.) The data were collected from electronic health records (EHRs) in a TriNetX Real-World database provided by a global health research network; with healthcare organizations spanning 17 countries, this system deploys a linked and continually updated global health research network representing over 300 million patients.

In the present work, we conducted a retrospective analysis of a large international COVID-19 cohort comprising 68,466 cases. Sub-cohorts were disaggregated by age (15+ years) and sex before statistical analyses were performed. The incidence data are presented in age steps of five years, from 5 to 86+ years for both sexes. Because of the expected estradiol hormone status, two subgroups of the female cohort were analyzed: we defined 15–49 years as the younger (pre-menopausal) group and 50+ for the older (post-menopausal) group. The group comparison was calculated for COVID-19–positive patients with supplemental estradiol versus subjects without estradiol in the respective age strata.

### Real-World Evidence Data

Using real-world evidence (RWE) data affords several advantages over more traditional approaches. Prospective, placebo-controlled, randomized, double-blinded multicenter clinical trials are the gold standard of evidence generation in medicine; however, such efforts are often slow and expensive. Furthermore, classical trials generally represent artificial situations with well-selected—and thus non-representative—pools of patients (e.g., often young men with few to no co-morbidities). Some of these issues can be alleviated by turning to RWE gleaned from EHR–derived data. Despite all this, one disadvantage of retrospective, implicit recruitments is the missing randomized placebo arm of the statistical study. Thus, generating suitable control cohorts is one of the most important aspects of an RWE-based study. One significant advantage of RWE is the large number of patients who can be recruited—this, in turn, yields relatively tighter confidence intervals and allows for cohort balancing or patient matching. In non-RWE studies, a confounder analysis can be performed, but doing so is made more difficult because of the relatively small number of patients enrolled.

### TriNetX

The TriNetX platform ensures data quality control by processes and procedures triggered in response to questions about the data provided. TriNetX provides details on the data provenance and/or offers the necessary access to audit the processes. TriNetX also makes the data available for a third-party audit.

## Results

The consolidated standard of reporting trial (CONSORT) flow diagram, shown in Figure 1, illustrates the data extraction process from the TriNetX Real-World database. The full cohort size of n = 68,466 patients consists of 37,086 women and 29,609 men who are older than 15 years (database accessed on July 16, 2020) and who are SARS-CoV-2 infected. After excluding (i) boys and girls up to 14 years of age, (ii) all men, and (iii) those without gender information, a sub-cohort of 37,086 women was used for further analysis of sex hormone status.

**Figure 1:**
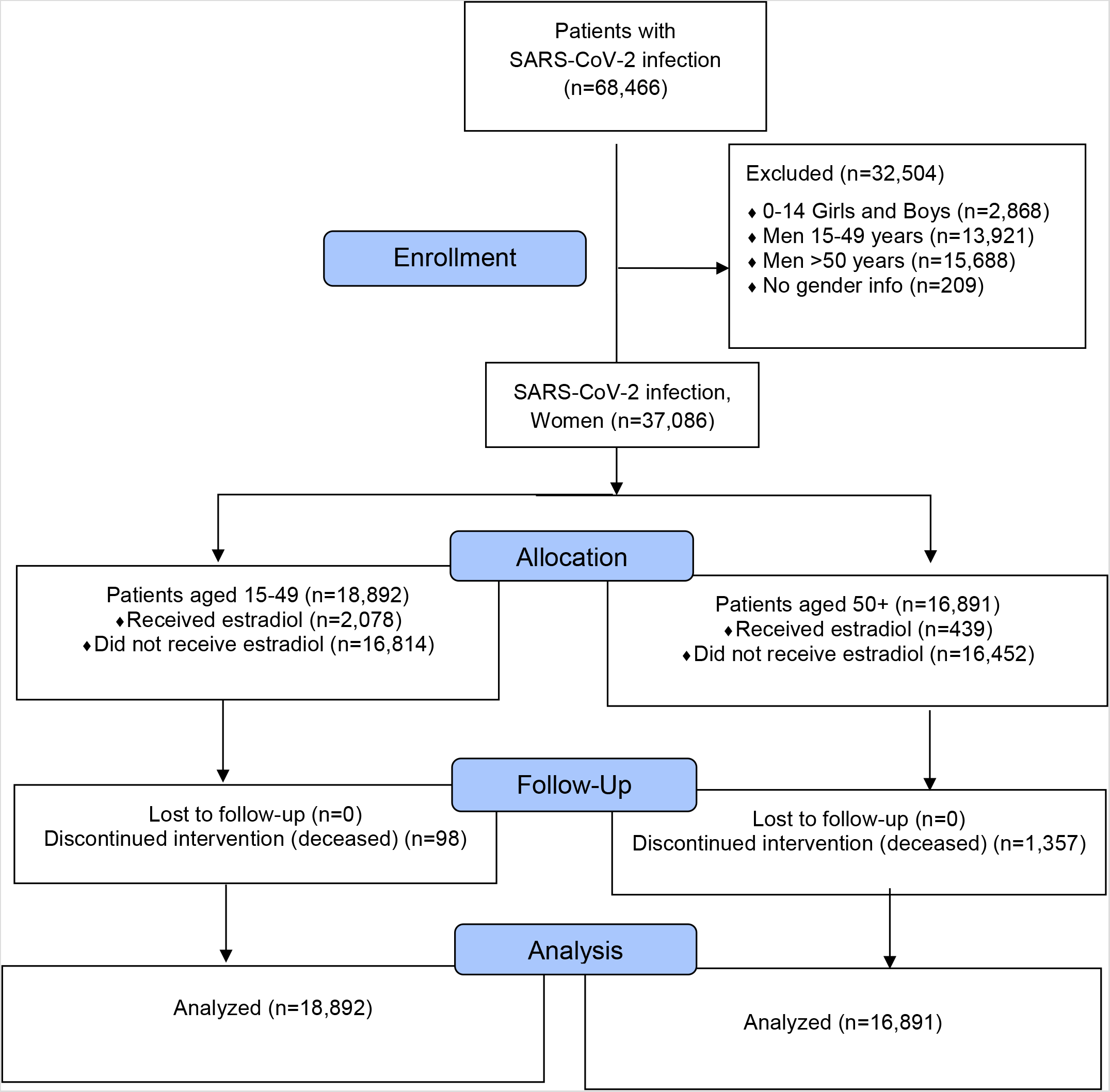
Our CONSORT flow diagram; major stages are indicated in the blue boxes.

Regarding gender medicine aspects and analyzing the data in terms of 5-year age strata, the incidence rates of women and men with known SARS-CoV-2 infection can be seen to differ by sex and age (Figure 2). The highest SARS-CoV-2 infection numbers among women, compared to age-matched men, is observed in women with what would be expected to be high levels of serum estradiol—i.e., between 15 and 49 years of age. The incidence of COVID-19 in women in this pre-menopausal age group is about +15% higher versus age-matched men (n = 21,229 women vs. n = 15,918 men). Starting with 60 and up to 85 years of age, the data show a relative decrease in frequency among women (n = 7,334 women vs. n = 7,171 men). In older women and andropause men (age strata 65 to 80), the statistical susceptibility to viral infection converges (see the closely-matched red and blue pairs of bars in Figure 2, in the range of 65–85-yrs old). Near the end of life, defined as 85+ years old, the incidence of SARS-CoV-2 infection is higher in women (n = 5,865) than in men (n = 5,406). At the opposite end of the age spectrum, note that all pre-adolescents (girls and boys) have similar risks of infection.

**Figure 2:**
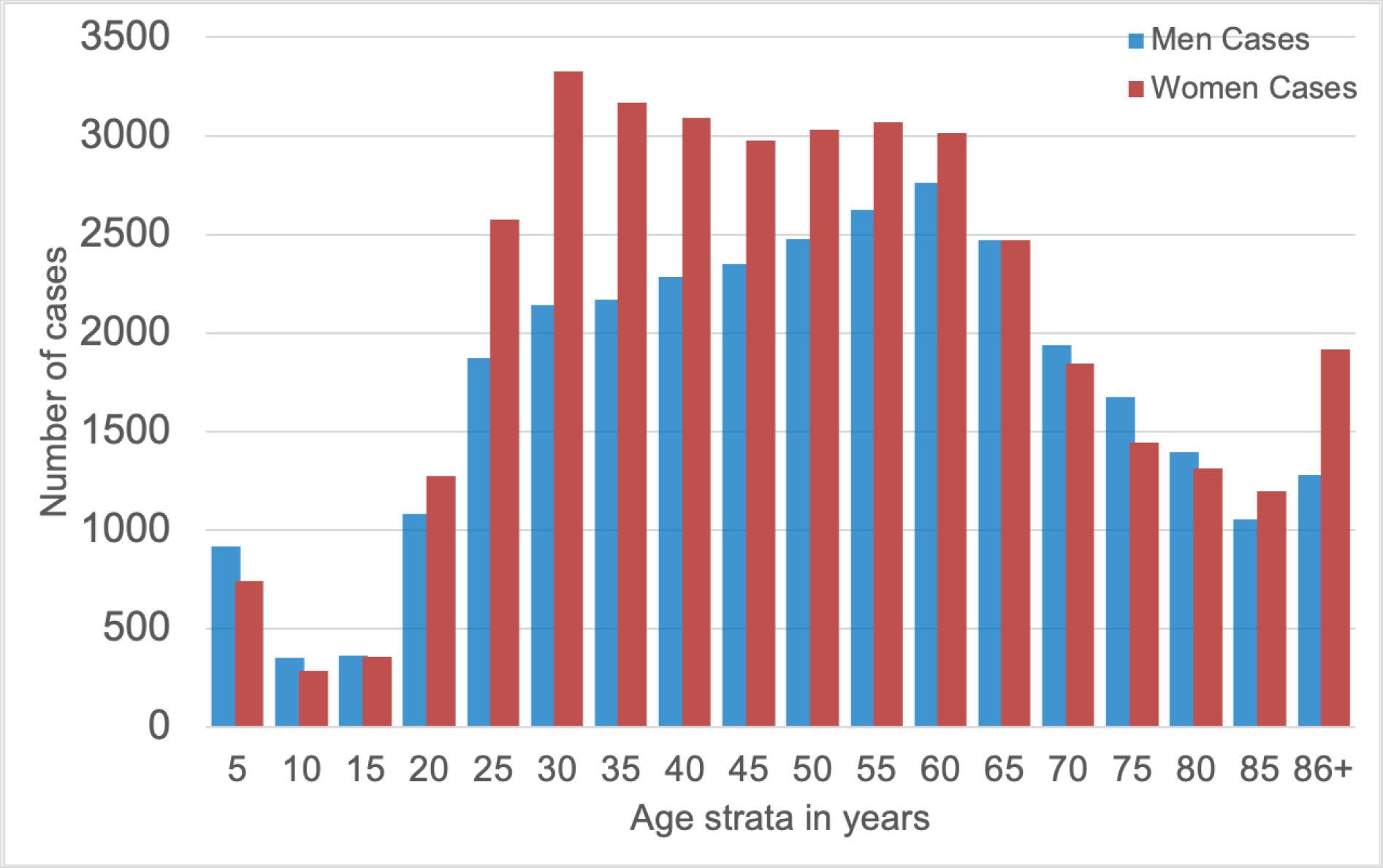
Absolute cases of COVID-19 in our cohort, disaggregated by women (red) and men (blue).

### Death Rate for the Full Cohort

The results of our data analysis for the death rate in women and men with COVID-19 are shown in Figure 3. Starting at age 45 years, the death rate increased for both men and women; men appear to be especially vulnerable to SARS-CoV-2 infection. The fatality rate is approximately +50% higher in men vs. women (Figure 3). For older men vs. women (those > 50 years of age), the calculated odds-ratio (OR) was 1.68, 95% CI [1.55, 1.81]. Correspondingly, a higher risk of death was associated with men; the hazard ratio (HR) was 2.7 [1.42, 5.08] with a lower survival probability for men than women. At a timepoint 200 days after diagnosis of SARS-CoV-2 infection, men were associated with a lower chance of survival than women (p< 0.0001).

**Figure 3:**
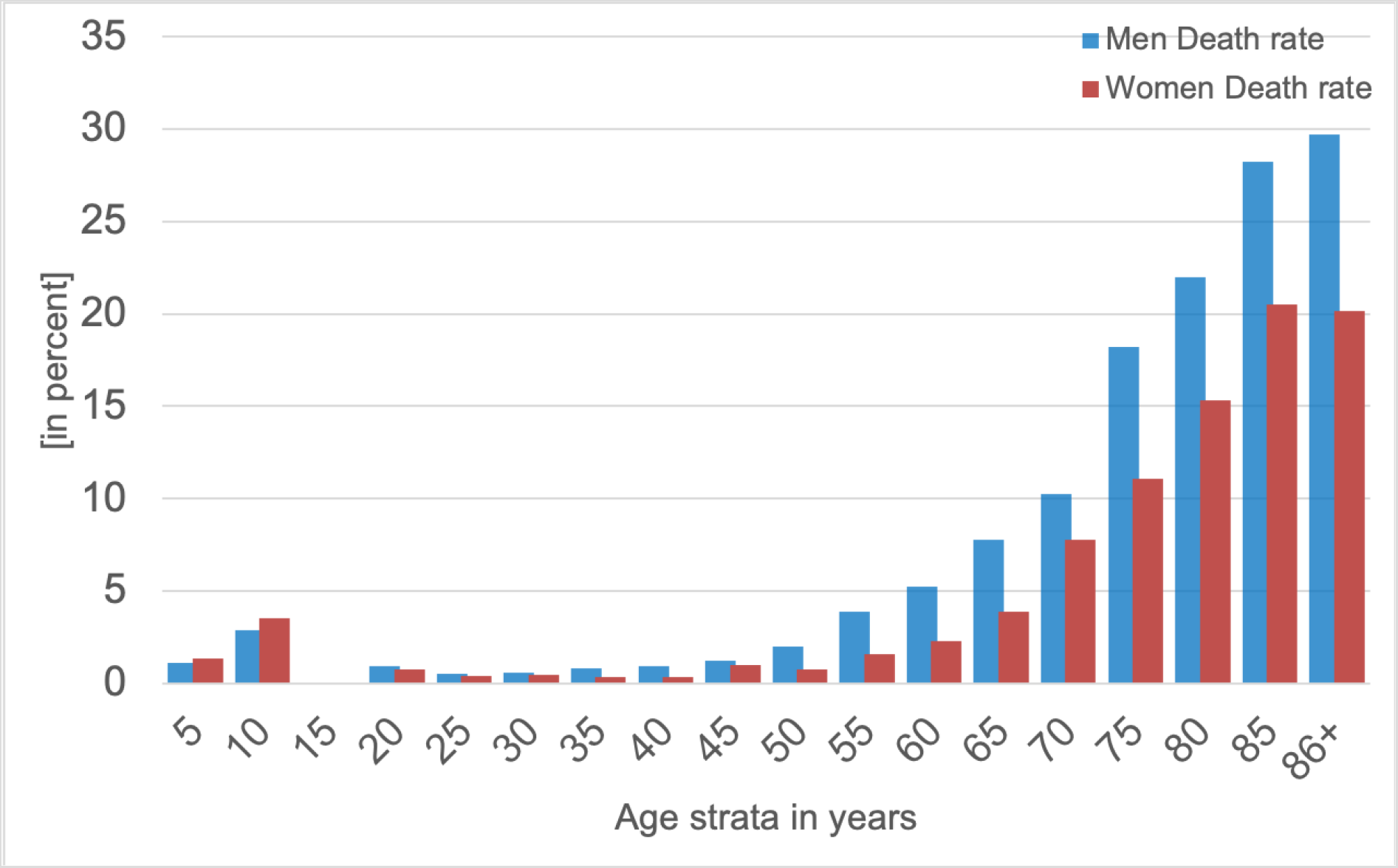
*A histogram of death rates for women (red) and men (blue) in five-year age strata:* Cumulatively, men were more vulnerable to SARS-CoV-2 infection, with a fatality rate in some strata that is roughly +50% higher than that in women.

**Figure 4:**
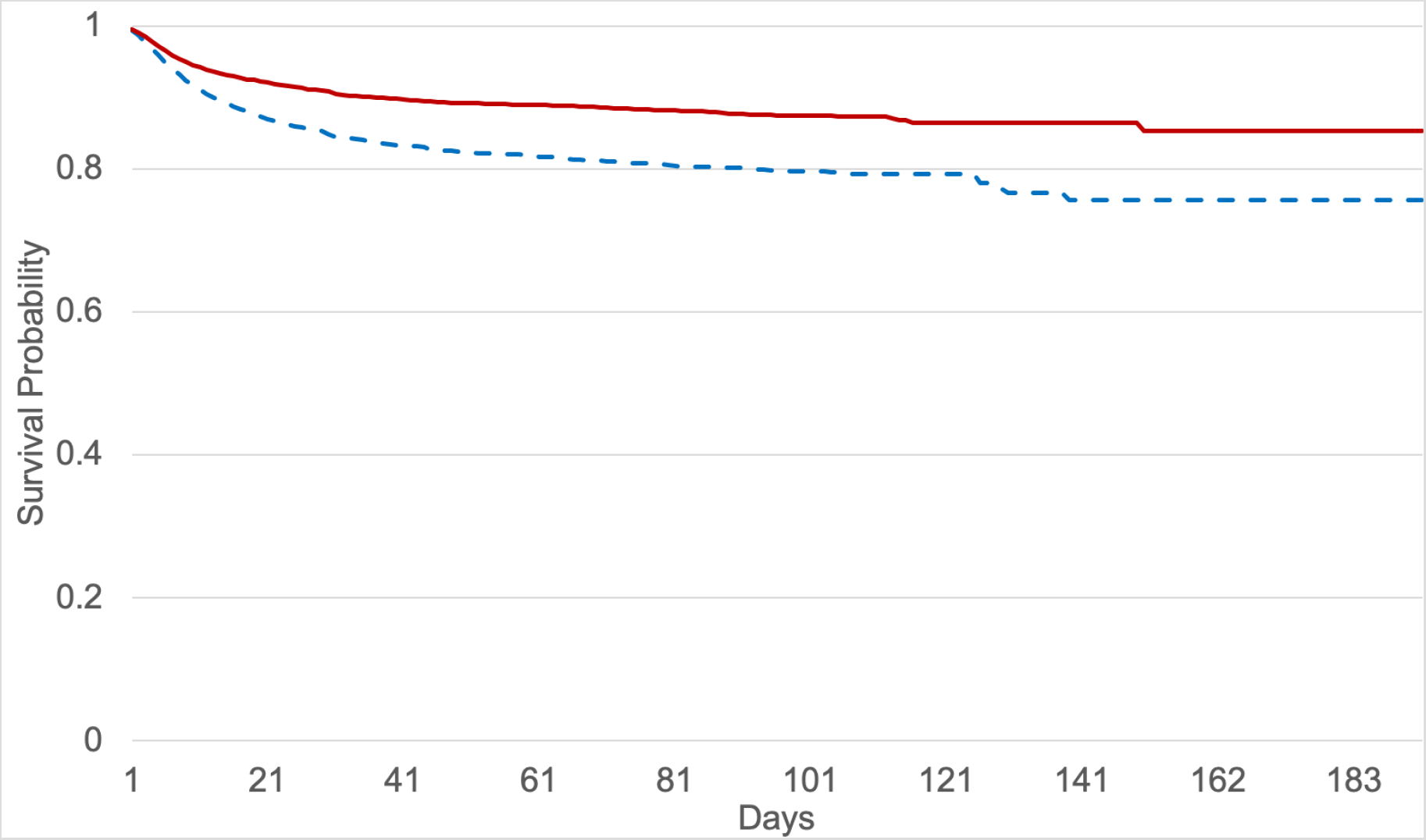
Kaplan-Meier survival curves for women (red) and men (blue, dashed), aged 50+ and with SARS-CoV-2 infection.

## Results for the Female Sub-cohort

### A Link between Exogenous Estradiol and Outcomes?

In order to assess the potential effects of exogenous female sex hormones in enhancing the survival likelihood for women with SARS-CoV-2 infection, we analyzed a sub-cohort of only women. A logistic regression analysis was performed for the combined outcome variable “death”. The outcome was calculated for n = 18,892 women, aged 15–49 years, who used oral contraceptives such as estradiol and ethinylestradiol (defined as a ‘user’ group), or without regular use of oral contraceptives (defined as ‘non-user’); we also included n = 16,891 peri- and post-menopausal women (50+), either with estradiol hormone usage (‘user’) or without regular usage of estradiol hormone (‘non-user’).

After matching the data of propensity scores for women aged 50+ with COVID-19, there was found to be a benefit for the estradiol hormone-user group vs. the non-user group, as regards an outcome of fatality. The OR calculated via logistic regression analysis for the combined outcome variable was 0.33 [0.18, 0.62] and the hazard ratio (HR) was 0.29 [0.11, 0.76] for the estradiol non-user vs. hormone-user group (Table 1). The average age across both groups was 64.2 years. The risk reduction for fatality from 6.6% (non-user) to 2.3% (user) was statistically significant (p< 0.0001).

**Table 1:**
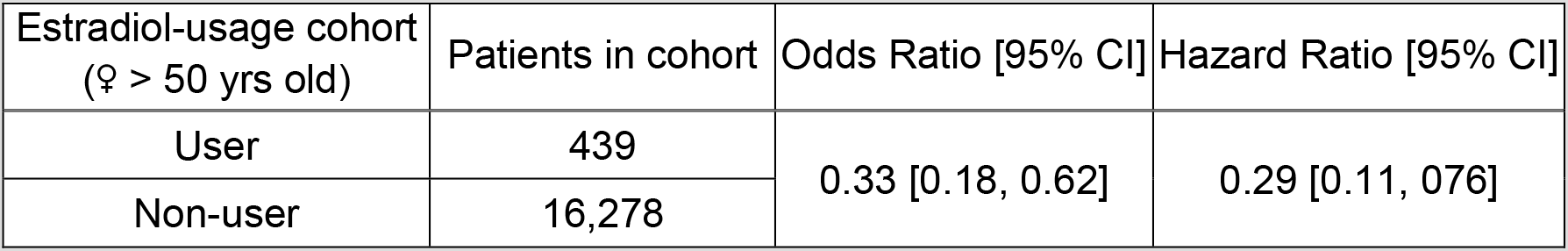
Cohort of estradiol user and non-user women aged 50+ Estradiol-usage cohort

| Estradiol-usage cohort<br>(♀ > 50 yrs old) | Patients in cohort | Odds Ratio [95% CI] | Hazard Ratio [95% CI] |
| --- | --- | --- | --- |
| User | 439 | 0.33 [0.18, 0.62] | 0.29 [0.11, 0.76] |
| Non-user | 16,278 |  |  |

The data for women 15–49 years of age, who were either documented users or non-users of oral contraception (estradiol, ethinylestradiol), were analyzed by logistic regression models. The effect was smaller than in the peri- and post-menopausal group (aged 50+). The OR calculated with logistic regression analysis for the combined outcome variable “death” was 1.0 [0.41, 2.4] for the non-user group vs. the oral contraception-user group (Table 2).

**Table 2:**
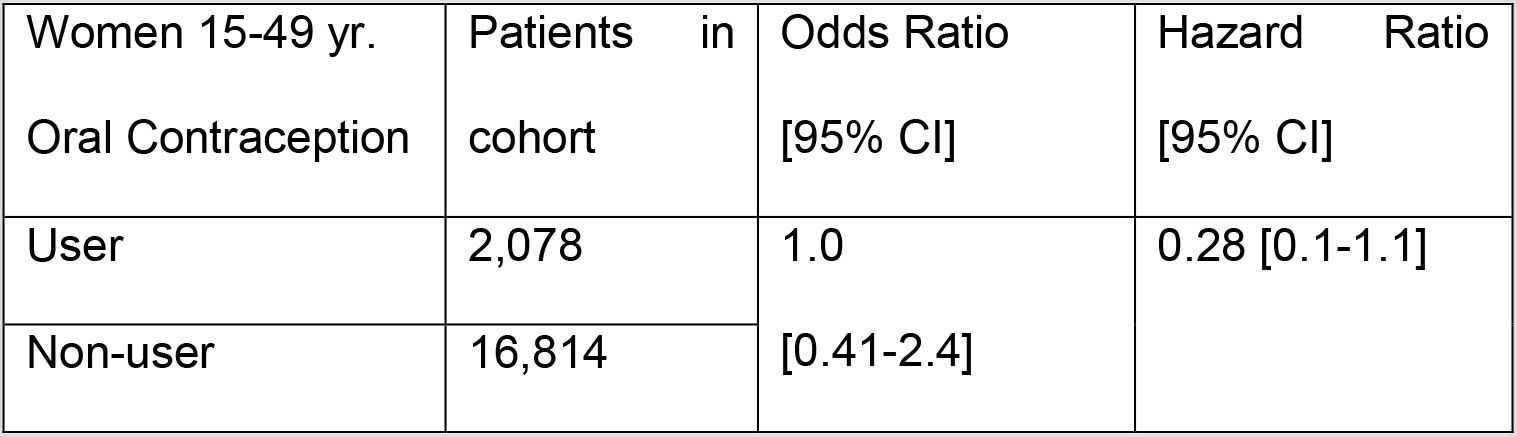
Cohort of oral contraception user and non-user women 15–49 years of age Women 15-49 yr.

| Women 15-49 yr.<br>Oral Contraception | Patients in<br>cohort | Odds Ratio<br>[95% CI] | Hazard Ratio<br>[95% CI] |
| --- | --- | --- | --- |
| User | 2,078 | 1.0<br>[0.41-2.4] | 0.28 [0.1-1.1] |
| Non-user | 16,814 |  |  |

### Survival Probability

Survival probabilities were calculated via Kaplan-Meier analyses, which revealed a significant difference (p< 0.0001) among women for the 50+ estradiol user group (n = 439) compared to the age-matched estradiol non-user group (n = 16,278). Considering the survival probability at 180 days after diagnosis of SARS-CoV-2 infection (index event), n = 10 women died in the estradiol user group and n = 1,072 died in the non-user group. Peri- and post-menopausal women, aged 50+, benefitted from estradiol hormone use: the 180-day survival probability for this cohort was 96.7%, compared to 84.9% for the non-user group (Figure 5).

**Figure 5:**
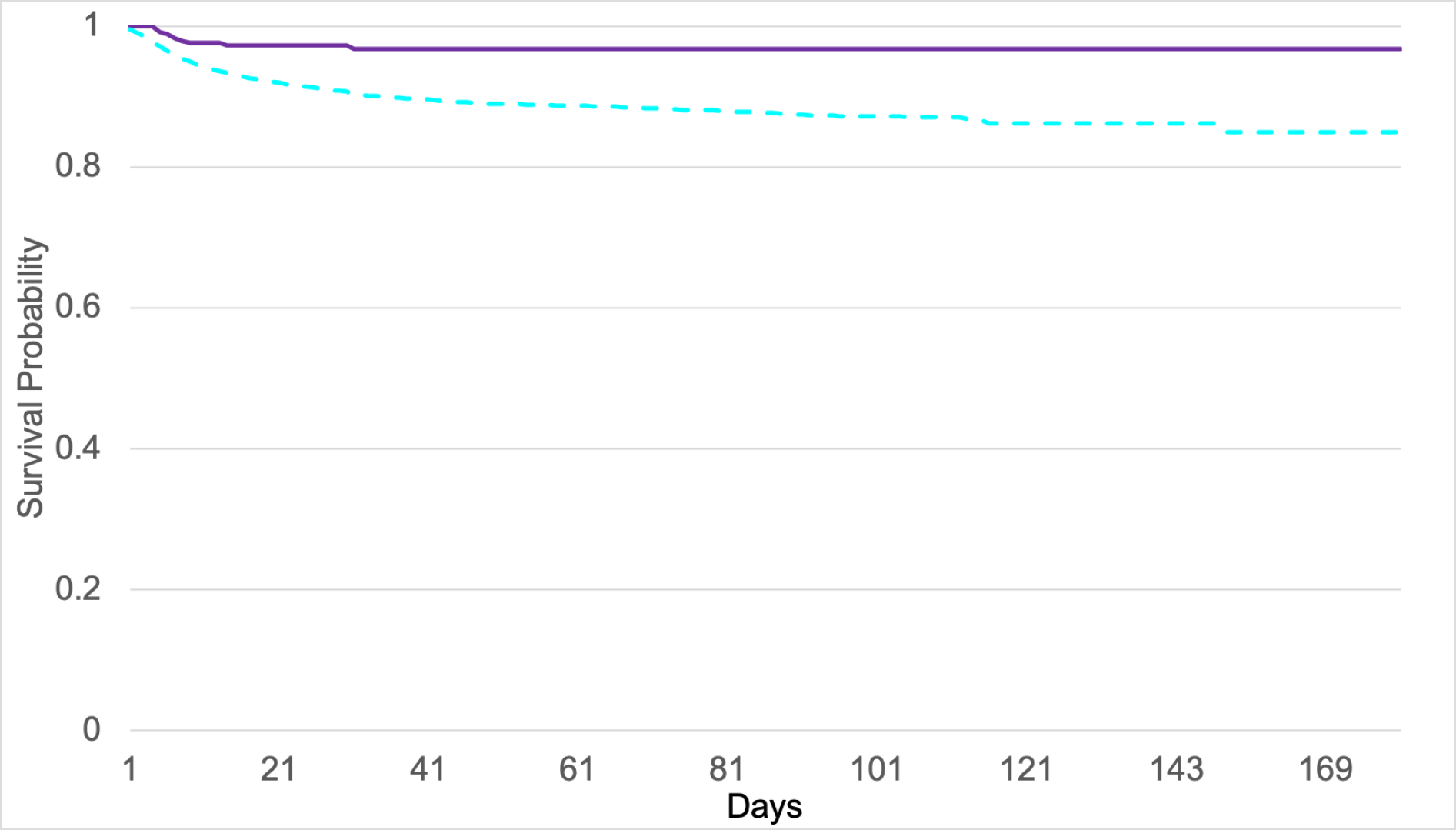
Kaplan-Meier survival curves: The survival probability of age 50+ women who were estradiol users (violet line) is shown, alongside non-users (blue dashed line).

## Discussion

This study focuses on the incidence and outcome of COVID-19 infections by considering an age- and sex-disaggregated data analysis. We identified a sex-specific distribution of COVID-19 incidence rates, with the highest frequencies being among premenopausal women in the 20–55-year age range. We also found a higher fatality rate of men compared to age-matched women, beginning at 50 years of age.

The patterns of incidence rates of women and men with SARS-CoV-2 infection, measured using 5-year age strata, differs from the trends in the number of deaths following the same age strata (Figure 3). This gender-based discrepancy might hint at different factors as contributing to the differential risks of infection and COVID-19 fatality among women versus men; note that these other factors may be either intrinsic (e.g., physiological/mechanistic) or extrinsic (e.g., smoking, lifestyle patterns or other circumstances) in nature. Gebhard et al. recently published the first sex-disaggregated data of confirmed COVID-19 cases and deaths, provided by Global Health 50%50 data tracker. Data from China and Europe were evaluated. Regardless country-specific demographics, case fatality in men resulted higher than in women through all age groups, and was more evident in the middle age (Gebhard, Regitz-Zagrosek, Neuhauser, Morgan, & Klein, 2020). Similarly, Klein et al. reported in the United States a significant male-female difference in COVID-19 cases, hospitalizations, and deaths (Klein et al., 2020).

The occurrence of most cases of SARS-CoV-2 infection within the reproductive ages—i.e., before age ∼50 (see Fig 2 and the dip towards the right-hand side)—hints at an association with relatively higher estrogen levels. General features of the datasets used in our large international COVID-19 cohort, comprising n = 68,466 cases, are consistent with the public data for the incidence of coronavirus infection and the fatality rate for COVID-19 disease in Germany, as presented by the Robert-Koch Institute (RKI) website (Robert-Koch-Institut, 2020). In our study the incidence of coronavirus infection in young women is about +15% higher than age-matched men; however, the fatality rate is about +50% higher in men than women.

The data in our present study indicate that pre-menopausal women are disproportionately more infected with coronavirus than men in the same age brackets, but they do not become as seriously ill, as evidenced by lower fatality rates; we believe this to be an interesting observation for sex and gender medicine experts.

Gender-specific epidemiological differences, with pathophysiological bases, have been published in the recent past, pointing to potential mechanistic roles for the sex hormones. However, X-chromosomal location of the ACE2- and the AT2-receptor genes and the downregulation of the pleiotropic cytokine interleukin 6 (IL-6) by estrogens, and its up- regulation by androgens, should also be considered other biochemical/physiological factors to which gender-correlated discrepancies might be attributed (Dorak & Karpuzoglu, 2012).

Figure 6 schematizes the potential downstream influence of 17β-estradiol on the reninangiotensin-aldosterone system (RAAS), including protective effects for target organs and tissues such as those of the lung, kidney, heart, cardiovascular system, adipose tissue, gut, testes, and the central nervous system. We emphasize that this diagram offers a tentative hypothesis for the mechanistic pathways that could be most salient in SARS-CoV-2 infections of pre-menopausal women with high estradiol levels. A central molecular component here is human ACE2, which is known to interact strongly with the SARS-CoV-2 viral spike protein (Turner, Hiscox, & Hooper, 2004) and, together with the “transmembrane protease, serine 2” (TMPRSS2) enzyme, play an essential role in viral entry into host cells (Hoffmann et al., 2020).

**Figure 6:**
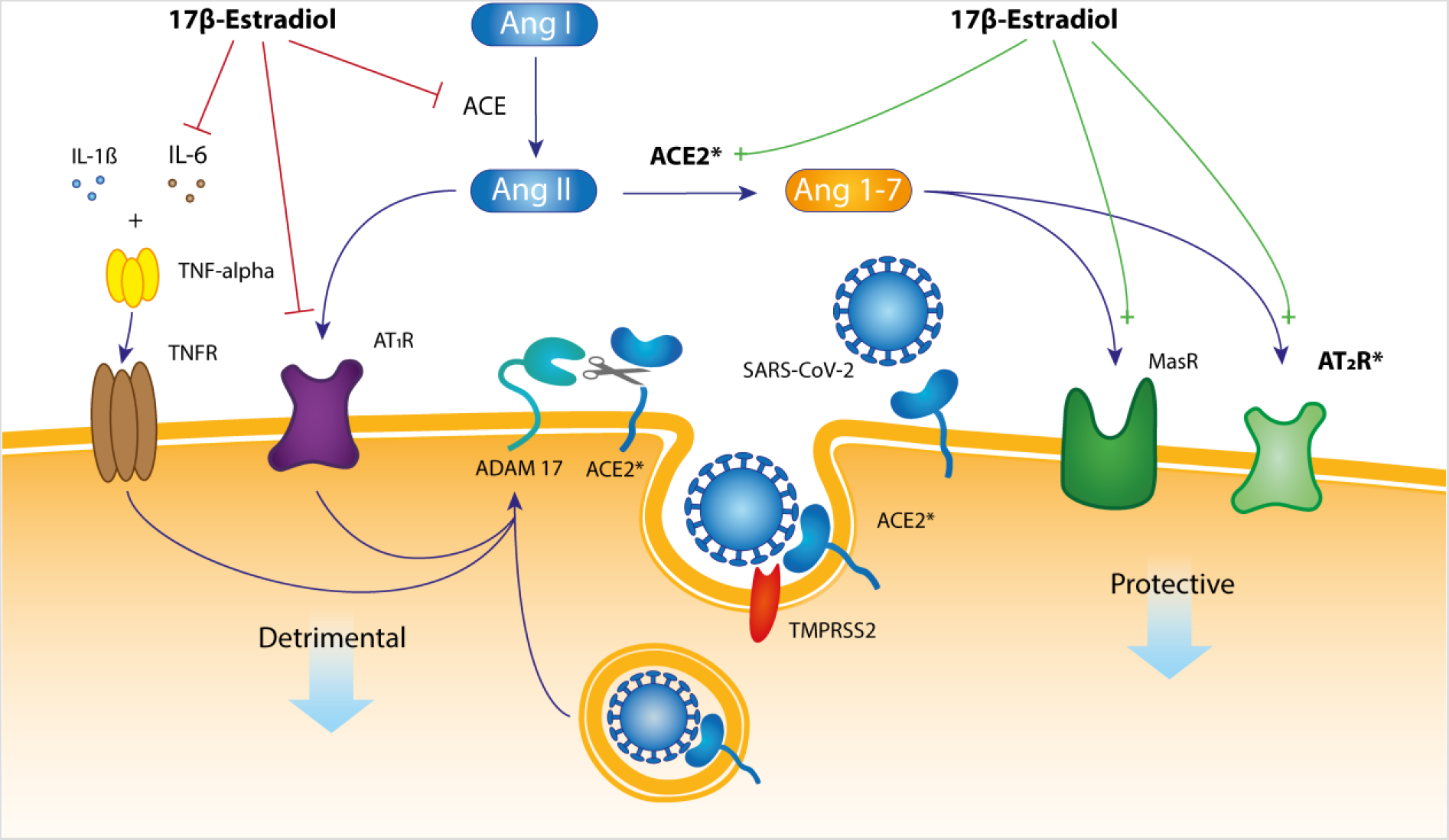
A hypothetical mechanistic pathway for the presumably protective role of 17βestradiol in SARS-CoV-2 infection. The membrane-tethered ACE2 protein has an amino-terminal catalytic domain (a peptidase) that faces the extracellular space.

In terms of molecular mechanisms, note that 17β-estradiol acts to block the IL-6 cytokine pathway and downregulate the angiotensin II (Ang II)–angiotensin II receptor type 1 (AT1R) signaling pathway. One route by which it does so is by inhibiting the activity of angiotensinconverting enzyme (ACE), as schematized in Figure 6. Also, as mentioned in the Introduction, the classical ACE–Ang II–AT1R regulatory axis and the ACE2–Ang 1–7–MasR/AT2R signaling pathway counter-regulate one another (Figure 6), and this is another way in which 17βestradiol can act: activation of the Mas receptor or AT2R activation via estradiol binding has a protective effect on target cells. Activation of the MasR and AT2R receptors protect target cells and organs by virtue of anti-fibrotic, anti-oxidant, anti-hypertrophic, and vasodilatory effects (Seeland & Regitz-Zagrosek, 2012). The 17β-estradiol hormone increases expression levels and activity of ACE2 in adipose tissue, kidneys, and myocardium (La Vignera et al., 2020).

Yet another potential molecular mechanism for estradiol’s effects stems from ACE2 and AT2R both being located on the X chromosome. TMPRSS2 (transmembrane protease serine subtype 2) expression is increased by estrogen receptor alpha (ERalpha) agonists such as estrogens (Janele et al. 2006) and, as diagramed in Fig 6, this protease is also involved in cellular uptake of SARS-CoV-2 virions. These systems also feature feedback: ACE2-mediated cell protection is lost following endocytosis of ACE2/SARS-CoV-2 (indeed, depletion of cell-surface ACE2 is a critical pathological outcome of SARS-CoV-2 infection), and this ACE2 shedding is mediated by three possible pathways that each elevate the activity of the metalloprotease known as ADAM-17. Moreover, ADAM-17 activity is followed by soluble tumor necrosis factor-α (TNF-alpha) release into the extracellular space (not shown explicitly in Fig 6). 17β-estradiol attenuates this latter pathway by inhibiting IL-6 activity (Fig 6). In conclusion, the levels of cell-surface–exposed ACE2 generally will be higher in women because of both (i) the balance between the effects of 17β-estradiol (activating and repressing) on the multiple pathways shown in Fig 6, and (ii) by virtue of the X chromosomal location of the ACE2 gene. Thus, while pre-menopausal women might be at a higher risk of initial SARSCoV-2 infection, they are more protected from a severe course of the disease.

Extending the hypothetical model and molecular factors described above, the relative dominance of the MasR- and AT2R-based receptor signaling pathways in pre-menopausal women has positive (beneficial) impacts upon downstream organ function, and it could represent a significant basis for the sex differences in SARS-CoV-2 infection; pre-menopausal women are more protected from cardiovascular, lung, and kidney diseases than their age-matched male peers. Recently, it has been shown that the estrogen-mediated up-regulation of the Mas-receptor contributes to the prevention of acute lung injury and affects endothelial barrier stabilization (Erfinanda et al., 2020). A relative protection of females over males has also been observed in other studies with experimental animal models of acute lung injury (Carey et al., 2007); intriguingly, this protection was lost in ovariectomized mice, but restored upon estrogen replacement (Speyer et al., 2005). The 17β-estradiol molecule attenuates lung vascular permeability and edema (Breithaupt-Faloppa et al., 2020), and estrogens reduce the pulmonary vasoconstriction during hypoxia by increasing levels of prostacyclin and nitric oxide (NO) (Breithaupt-Faloppa et al., 2020). Moreover, in experimental mice models, active ACE2 alleviates pulmonary injury and vascular damage (Imai et al., 2005; Kuba et al., 2005). These animal model experiments point to mechanisms that underlie the organ-protective effects of 17β-estradiol, such as anti-fibrotic, anti-oxidant, anti-hypertrophic, and vascular dilation effects.

We assume that premenopausal women have higher basal ACE2 serum levels than age-matched men—a scenario that could translate into a higher risk of SARS-CoV-2 infection in women. Higher basal ACE2 levels could be due to the effects of estradiol on RAAS (Figure 6) and/or the fact that the ACE2 and AT2R genes map to the X chromosome. In genetic terms, this sex-linkage would allow women to be heterozygous and differently assorted compared to men, who are hemizygous (Gemmati et al., 2020). The second X chromosome is not inactivated in approximately 15 % of genes consistently escape from this inactivation and another 15 % of genes vary between individuals or tissues in whether they are subject to, or escape from, inactivation (Balaton, Cotton, & Brown, 2015). This may account for some of the differences that are seen between men and women (sexual dimorphism) and could be a reason for higher expression levels of ACE2- and ATIIR proteins in women (i.e., a gene dosage effect).

Another pertinent mechanistic pathway, potentially triggered or modulated by the types and levels of sex hormones, could be immunological—namely, gender-specific differences in the immune response to SARS-CoV-2 infection. A recent study has linked higher mortality among men to a “cytokine storm”, which in turn closely relates to the severity of symptoms such as pulmonary edema, fibrosis and other deleterious downstream effects associated with acute lung injury (Li et al., 2020). An individual’s immune response to viral infections can vary with fluctuations in sex hormone concentrations, including steroid hormones such as estrogens, progesterone, and testosterone. Each immune cell type is influenced by all three of these major sex hormones, albeit with different and partly opposing effects. From a biomedical perspective of sex and gender, interleukin-6 (IL-6) is particularly interesting. IL-6 is a cytokine with both anti- and pro-inflammatory effects. It can be produced by almost all stromal and immune system cells (e.g., monocytes, lymphocytes, macrophages, endothelial cells, mast cells, dendritic cells), and it is believed to play a central role in precipitating the cytokine storms that, in turn, yield such severe symptoms from a SARS-CoV-2 infection (McGonagle, Sharif, O’Regan, & Bridgewood, 2020). Notably, the endogenous female sex hormone 17β-estradiol blocks the IL-6 cytokine pathway, while the male sex hormones (the androgens) increase IL-6 production (Dorak & Karpuzoglu, 2012).

Given the general, evidence-based model that we have presented above—namely, that the female sex hormone estradiol is closely coupled to a broad array of downstream physiological effects, including those related to SARS-CoV-2 infection—the second part of our retrospective data analysis examines the outcome (required ventilation and death) of pre- and postmenopausal women with SARS-CoV-2 infection, including as a function of whether or not exogenous estradiol is in use. In performing this analysis, we again employed the large, international COVID-19 cohort developed in this study. To our knowledge, this is the first report that has evaluated the impact of exogenous estradiol sex hormonal use, either for contraceptive purposes or for postmenopausal symptoms, on COVID-19 fatality.

Among post-menopausal women, we observed a significant difference in the rates of death between women with regular estradiol use (user group) and those without estradiol sex hormone intake (non-user group). Furthermore, our data analysis included distinct sub-groups of young women (15–49 years of age) with and without oral contraceptives. The dominant form of estrogen used for “the pill” is ethinylestradiol. The magnitude of the protective effect of the usage of oral contraception, compared to the non-users sub-population, is smaller among younger versus older women; this may be the case because endogenous estradiol levels are typically already higher in younger women than for post-menopausal women, thus drowning-out any differences between user/not-user groups. Moreover, the level of exogenous hormone intake for purposes of contraception is generally less than that used for post-menopausal estradiol hormone therapy.

Note that other current trials are testing the effect of sex hormones (estrogen and testosterone) on COVID-19 outcomes. A brief, seven-day course of estradiol, delivered via a transdermal patch, could be a safe approach to reduce symptom severity in adult men and in older women, when administered prior to intubation (https://ClinicalTrials.gov; Identifier: NCT04359329). The time-course for estradiol treatment may need to be evaluated for its positive effects on lung protection, and whether it could be an effective therapeutic approach not only for women but also for men with COVID-19. This type of question cannot be answered solely by the data presented here, and instead addressing this question is a key topic for further research.

Based on the main findings of our present study, we believe there are no concerns for continuing the use of sex hormones that contain estradiol prior to SARS-CoV-2 infection. Even though it can be seen in the data that the risk of infection is higher in pre-menopausal women with higher endogenous estradiol levels, compared to either men of the same age strata or to post-menopausal women, the clinical course of COVID-19 disease, and the ultimate mortality rate, is lower in women with higher estradiol levels. Higher survival probabilities are particularly evident in post-menopausal women who are infected with SARS-CoV-2 and who regularly use exogenous estradiol.

## Conclusions

Pre-menopausal women are at a relatively high risk for SARS-CoV-2 infection, but the survival probability in this below-50 age range is significantly higher in women than in men. A chief finding of this study is the strong positive effect of regular estradiol hormone therapy on the survival rates of post-menopausal women.

## Data Availability

We utilized the TriNetX platform. TriNetX provides details on the data provenance and/or offers the necessary access to audit the processes. TriNetX also makes the data available for a third-party audit.

## List of abbreviations

COVID-19: Coronavirus Disease 2019
ACE: Angiotensin-converting enzyme
OR / HR / CI: Odds Ratio / Hazard Ratio / Confidence Interval
SARS-CoV-2: Severe Acute Respiratory Syndrome-Coronavirus-2
AT2R: Angiotensin II receptor Type 2
RAAS: Renin angiotensin aldosterone system
IL-6: Interleukin-6
Ang1–7: Angiotensin 1–7
EHR: Electronic Health Records
RWE: Real-world evidence

## Declarations

### Ethics approval and consent to participate

Not applicable.

### Consent for publication

Not applicable

### Availability of data and materials

The data that support the findings of this study are available from TriNetX. While restrictions apply to the availability of these data, which were used under license for the current study and are not publicly available, the data are available from the authors upon reasonable request and with the permission of TriNetX.

### Authors’ contributions

US analyzed and interpreted data, wrote the first version of the manuscript, designed Figures about mechanistic pathways.

MS analyzed and interpreted data.

FC analyzed and interpreted data.

CM was involved in interpretation of the data and helped write the manuscript.

PEB was involved in interpretation of the data and helped write the manuscript. MH analyzed and interpreted data.

RP conceived the study, matched cohorts, and performed the analysis. SP conceived the study, performed the analysis, and designed Figures. All authors read and approved the final version of the manuscript.

## Acknowledgments

Not applicable

## Authors’ information

Not applicable.

